# Prolonged infusion versus intermittent infusion dosing of beta-lactam antibiotics in critically ill patients with sepsis: a protocol for a systematic review and meta-analysis of randomised controlled trials

**DOI:** 10.1101/2023.05.15.23289889

**Authors:** Mohd H. Abdul-Aziz, Naomi E Hammond, Stephen J. Brett, Menino O. Cotta, Jan J. De Waele, Gian Luca Di Tanna, Joel M. Dulhunty, Hatem Elkady, Lars Eriksson, M. Shahnaz Hasan, Jeffrey Lipman, Giacomo Monti, John Myburgh, Emmanuel Novy, Dorrilyn Rajbhandari, Claire Roger, Joseph Alvin Santos, Fredrik Sjövall, Irene Zaghi, Alberto Zangrillo, Anthony Delaney, Jason A. Roberts

## Abstract

**Introduction:** *In vitro* and *in vivo* pharmacokinetic/pharmacodynamic data describe improved activity of beta-lactam antibiotics when administered by prolonged infusion compared with standard intermittent infusion. There remains insufficient robust clinical trial data to support a widespread practice change. Patients with sepsis and septic shock are a population in whom prolonged infusion of beta-lactam antibiotics may improve survival. Two large multicentre randomised controlled trials (RCTs) comparing prolonged versus intermittent infusion of beta-lactam antibiotics in critically ill patients with sepsis or septic shock are due for completion in 2023. With existing RCT evidence, this systematic review and meta-analysis will include these new data to measure the clinical benefits of prolonged beta-lactam infusion in critically ill patients with sepsis.

**Methods and analysis:** This protocol has been prepared according to the Preferred Reporting Items for Systematic Review and Meta-Analysis Protocols (PRISMA) statement. This systematic review and meta-analysis will include RCTs that compare prolonged infusion with intermittent infusion of beta-lactam antibiotics in critically ill adult patients with sepsis. Medline (via PubMed), CINAHL, EMBASE, Cochrane Central Register of Controlled Trials (CENTRAL), and other clinical trials registries will be searched to identify eligible RCTs for review. Two reviewers will perform the study selection and extraction processes with disagreements resolved by discussion or referral to a third reviewer if needed. The Cochrane Collaboration’s Risk-of-Bias Tool for Randomised Trials version 2 (RoB 2) will be used to evaluate the quality of included studies. The Grading of Recommendations Assessment, Development, and Evaluation (GRADE) approach will be used to evaluate the overall quality of evidence for each outcome measures The *a priori* primary outcome is all-cause 90-day mortality. Secondary outcomes include intensive care unit (ICU) mortality, ICU length of stay, clinical cure, microbiological cure, and the development of adverse events. Bayesian random-effects meta-analyses will be conducted, with frequentist analyses planned for sensitivity analysis.

**Ethics and dissemination:** Human research ethics approval is not required as the study involves the use of existing collections of data that are de-identified. It is expected that findings will be presented at national and international intensive care and infectious diseases meetings, and will be submitted to a peer-reviewed journal for publication.

PROSPERO Registration Number: CRD42023399434

## Introduction

### Rationale

Patients who develop sepsis and septic shock often require treatment in an intensive care unit (ICU) and face high morbidity and mortality rates (1). There is an urgent need to define strategies and interventions that can improve morbidity and mortality, as well as to reduce significant healthcare resource utilisation associated with sepsis management. Source control of the infection, along with early and appropriate antibiotic administration are central to the management of critically ill patients with sepsis (2). However, administering appropriate antibiotic therapy can be challenging in the ICU for a variety of reasons. Physiological changes can occur from pharmacological interventions such as the administration of fluid therapy, and the natural course of sepsis may also alter antibiotic pharmacokinetics in critically ill patients (3). In addition, pathogens isolated in the ICU are commonly less susceptible to common antibiotics than those in other environments (4). Conventional antibiotic dosing rarely considers these issues and therefore, has a higher likelihood to fail in this patient population (5-7).

The beta-lactam class of antibiotics are widely used to treat patients with sepsis or septic shock in the ICU due to their wide spectrum of antibiotic activity and favourable safety profile (8, 9). Beta-lactam antibiotics display “time-dependent” bactericidal activity, which is optimal when the duration of time (T) that the free drug concentration remains at least 40 – 70% of the time above the minimum inhibitory concentration (MIC) during a dosing interval (fT_>MIC_ or 40 – 70% fT_>MIC_) (10). However, recent data suggest that patients may benefit from higher (e.g., 2 – 5 x MIC) (11) and longer (e.g., 100% fT_>MIC_) (7, 12) beta-lactam antibiotic exposures than those described in earlier pre-clinical infection models (13). Therefore, administration via prolonged infusion (infusion duration ≥2 hours or greater) is theoretically advantageous compared to standard intermittent infusion, which is characterised by high peaks followed by low concentrations for longer periods of the dosing interval.

*In vitro* and *in vivo* pharmacokinetic/pharmacodynamic data show that prolonged infusions more consistently achieve effective beta-lactam antibiotic exposure associated with maximal bacterial killing than intermittent infusion (14, 15). Clinical studies reporting patient outcomes with prolonged infusion of beta-lactam antibiotics have varied, ranging from no significant effect (16-26), to significant improvements in patient mortality (27, 28), clinical cure (29, 30), microbiological cure (31), length of ICU and/or hospital stay (32, 33), and duration of mechanical ventilation (29). Most meta-analyses have included heterogenous patient populations (34-38), including those in whom a difference in effect between prolonged and intermittent infusions is unlikely (e.g., non-critically ill patients), or studies with other important methodological shortcomings (39).

Two large multicentre randomised controlled trials (RCTs) comparing prolonged versus intermittent infusion of beta-lactam antibiotics in critically ill patients with sepsis or septic shock are due to be published in 2023: (1) the Beta-Lactam InfusioN Group (BLING) III Study, which aims to recruit 7000 ICU patients across Australia, Belgium, France, Malaysia, New Zealand, Sweden and the United Kingdom (40), and (2) the continuous infusion versus intermittent administration of MERopenem in criticallY ill patients (MERCY) Study, which aims to recruit 300 ICU patients across Croatia and Italy (41). To provide context for clinicians to interpret the results of these studies in light of the larger body of evidence, we plan to perform a systematic review and meta-analysis to assess whether in critically ill patients with sepsis or septic shock, a prolonged infusion of beta-lactam antibiotic compared to standard intermittent bolus dosing is associated with reduced 90-day all-cause mortality, as well as assessing the effect on other prespecified secondary outcomes.

### Objective

The primary objective is to determine whether prolonged infusion of a beta-lactam antibiotic is associated with improved all-cause 90-day mortality when compared with intermittent infusion in critically ill adult patients with sepsis or septic shock. Key secondary outcomes will include ICU mortality, ICU length of stay, clinical cure, microbiological cure, and adverse events.

### Methods and analysis

This systematic review and meta-analysis of RCTs comparing prolonged versus intermittent beta-lactam antibiotic infusion in critically ill adult patients with sepsis or septic shock will follow reporting recommendations of the Cochrane Collaboration and the Preferred Reporting Items for Systematic Reviews and Meta-Analyses (PRISMA) statements (42). This systematic review has been registered on International Prospective Register of Systematic Reviews (PROSPERO) CRD42023399434.

### Eligibility criteria

#### Inclusion criteria

RCTs comparing prolonged versus intermittent infusion of one or more beta-lactam antibiotics, which meet the following criteria will be included:

- *Population* Critically ill adult (≥18 years old) patients with sepsis or septic shock receiving care in the ICU. All conventional and current definitions of sepsis and septic shock at the time of patient recruitment will be accepted (43-45). A study is determined to have been conducted in a critically ill population if the manuscript reported any of the following:
  1. the patients were recruited in an ICU, or
  2. the inclusion criteria described were such that the patients would normally be managed in an ICU (e.g., patients receiving invasive mechanical ventilation), or
  3. the patients were suffering from a condition that usually requires care in an ICU (e.g., severe burns of >40% total body surface area), or
  4. the patients had an average ICU length of stay of ≥2 days, or
  5. a majority of the patients received a therapy that is delivered in the ICU (e.g., invasive mechanical ventilation), or
  6. a severity of illness score which reflected a critically ill population.

ICU may include a general ICU or complex of ICUs (medical, surgical, or mixed) capable of providing close monitoring and support for critically ill patients with life-threatening conditions.

- *Intervention* Prolonged infusion of a beta-lactam antibiotic, where “prolonged infusion” is defined as either:
  - Extended infusion: intravenous drug administration for ≥2 hours during a dosing interval OR
  - Continuous infusion: constant intravenous drug administration either as a sequential 6-hour, 8-hour, 12-hour or 24-hour infusion.

- *Comparator* Intermittent infusion of a beta-lactam antibiotic where “intermittent infusion” is defined as administration of an intravenous drug infusion for <2 hours.

- *Outcomes* Studies that report or are able to provide any of the *a priori* primary or secondary outcomes specified in this systematic review and meta-analysis.

#### Exclusion criteria

The following studies will be excluded:

- Retrospective cohort studies
- Trials of patients not meeting the criteria for sepsis or septic shock.

### Search strategy

Medline (via PubMed), CINAHL, EMBASE, Cochrane Central Register of Controlled Trials (CENTRAL), pre-print servers (medRxiv and OSF Preprints), and clinical trials registries will be searched to identify eligible trials to be included for review. The search will be performed with no restrictions on language, publication date or publication status. We will use a combination of keywords and search terms to identify RCTs in:

- “sepsis” or “septic shock” or “systemic inflammatory response syndrome” AND
- “beta-lactam” or “carbapenem” or “cephalosporin” or “monobactam” or “penicillin” AND
- “continuous infusion” or “extended infusion” or “prolonged infusion” AND
- “critically ill” or “intensive care unit”

The search terms for this review will be created by a research librarian in collaboration with content area experts. Additionally, we will manually check the reference lists of relevant primary and review articles, as well as contacting experts in the field, to identify additional RCTs that may be eligible for inclusion. Full details of the electronic search strategy are available in the appendix.

### Study records

#### Selection process

Study titles and abstracts from the search will be screened in a reference management system (Covidence systematic review software, Veritas Health Innovation, Melbourne, Australia). Duplicates and irrelevant studies will be excluded. Review of titles and abstracts will be independently undertaken by two reviewers. Reports identified by either reviewer that may potentially meet inclusion criteria will be obtained for full text review. Full text manuscripts of potentially eligible studies will be assessed by two reviewers independently, with disagreements resolved by consensus or resort to a third reviewer if required. The selection process will be documented and presented in a Preferred Reporting Items for Systematic review and Meta-Analyses (PRISMA) flow diagram.

#### Data collection

Data from included studies will be extracted using a standardised data collection form. Data extraction will be performed in duplicate and any disagreements will be resolved by discussion or, if required, by referral to a third reviewer. Attempts will be made to contact corresponding authors to obtain essential additional data. Access to aggregate level data for the BLING III (40) and MERCY (41) studies prior to publication has been agreed by the respective investigators and study management committees. Data from unpublished studies will not be made public without the express prior consent of the responsible parties.

The following data will be extracted:

- Study characteristics: first author, year of publication, study period, recruiting countries, number of patients enrolled.
- Participant characteristics: age, sex, severity of illness scores at baseline, renal replacement therapy at baseline, renal replacement therapy during study period, microbiological confirmed infection (i.e., culture-positive infection), distribution of isolated pathogens (Gram-negative versus Gram-positive organisms), and site of infection.
- Study intervention and comparator details: antibiotic, dosing regimen, and concomitant antibiotics.
- Outcomes: 90-day mortality (or closest time point before and beyond), ICU mortality, ICU and hospital length of stay, clinical cure and the definition used in the study, microbiological cure and the definition used in the study, and the number of adverse events.

### Outcomes

#### Primary outcome

The primary outcome is all-cause 90-day mortality. If 90-day mortality outcomes are not reported in a study, we will use the time closest to Day 90 (before and beyond).

#### Secondary outcomes

Where available, the following secondary outcomes will be reported:

- ICU mortality
- ICU length of stay as reported in the original study
- Clinical cure as defined in the original study
- Microbiological cure as defined in the original study
- Adverse events as defined in the original publication

### Risk of bias

The Cochrane Collaboration’s Risk-of-Bias Tool for Randomised Trials version 2 (RoB 2) will be used to evaluate the quality of included studies. The tool will evaluate all types of bias that can affect results of RCTs covering five domains including bias arising from the randomisation process, deviations from intended interventions, missing outcome data, measurement of the outcome and selection of the reported result. For each domain, studies will be judged to either have “low risk of bias”, “some concerns” or “high risk of bias”. A proposed judgement will be generated by an algorithm based on answers to the signalling questions of the tool. The risk of bias assessment will be performed by two independent assessors who were not involved in any of the included studies.

### Statistical analysis

The meta-analysis will be based on a Bayesian (primary approach) and a frequentist (secondary approach) framework. Random-effects meta-analyses will be carried out and pooled effect estimates will be reported as Risk Ratio (RR) with 95% confidence interval (CI) or credible interval (CrI) for binary outcomes, and as Mean Difference (MD) for continuous outcomes. When median and interquartile range or range are reported, mean and standard deviation will be estimated using the method described by Wan et al (46).

For the Bayesian analysis, pooled effect estimates and posterior probabilities that prolonged infusion of beta-lactam antibiotics is associated with better outcomes compared to intermittent infusion will be generated using: (a) vague priors for the effect and heterogeneity parameters in the main analysis, and (b) weakly-informative priors in the sensitivity analysis. Normally distributed priors will be used for the effect parameters logRR and MD (e.g., a vague prior for the logRR centered at mean of 0 with a standard deviation of 2 will be used for binary outcomes), while half-normal priors will be used for the heterogeneity parameter τ^2^ (e.g., a vague prior of 0.5).

Where applicable and appropriate, weakly informative priors for the heterogeneity parameter will be specified for different types of outcome measures (47, 48). For the frequentist analysis, the (a) Hartung-Knapp-Sidik-Jonkman method, and the (b) DerSimonian-Laird method will be employed to obtain an overall effect estimate for each outcome measure.

Quantitative heterogeneity will be assessed using τ^2^ and its 95% credible interval. The proportion of variation across studies due to heterogeneity rather than chance will be assessed using the I^2^ statistic. Presence of small-study effects will be assessed through regression-based Egger’s test and visual inspection of the contour-enhanced funnel plots.

Statistical analyses will be performed using Stata BE V17 for Windows (StataCorp LLC, College Station, TX) and the bayesmeta package in R (49).

#### Missing data

Data from “intention-to-treat” populations will be used in the analysis and an attempt to obtain missing outcome data from the original study authors will be made. There will be no imputing of values for missing data.

#### Sub-group analysis

Sub-group analyses for the primary outcome will be hypothesis generating. The following patient sub-groups will be analysed if enough baseline data are available:

- Meropenem vs. piperacillin/tazobactam. It is hypothesised that improvements in patient survival will be greater in patients receiving prolonged infusion of piperacillin/tazobactam as longer % fT_>MIC_ exposure is required for antibiotic efficacy when compared with meropenem.
- Culture-positive vs. culture-negative infections. Patients with microbiological confirmed infections who receive prolonged infusion of beta-lactam antibiotics is hypothesised to show greater improvements in patient survival when compared with intermittent infusion.
- Gram-positive vs. Gram-negative infections. Patients with Gram-negative infections who receive prolonged infusion of beta-lactam antibiotics is hypothesised to show greater improvements in patient survival when compared with intermittent infusion. Gram-negative microorganisms tend to have higher MICs and, in such infections, pharmacokinetic/pharmacodynamic data have consistently demonstrated that prolonged infusion of beta-lactam antibiotics is more likely to achieve higher % fT_>MIC_ exposures for maximal bacterial killing.
- Renal replacement therapy vs. non-renal replacement therapy. It is hypothesised that improvements in patient survival will be greater in patients who receive prolonged infusion of beta-lactam antibiotics who are not on renal replacement therapy support. Patients receiving renal replacement therapy are likely to have reduced drug clearance leading to higher and longer % fT_>MIC_ beta-lactam antibiotic exposures, regardless of which administration method is used.
- Lung infections vs. other infections. It is hypothesised that improvements in patient survival will be greater in patients with lung infections who receive prolonged infusion of beta-lactam antibiotics. An administration method that can enhance the beta-lactam antibiotic penetration into the interstitial fluid of the infected lung tissues (where the antibiotic-bacteria interactions occur) is likely to improve patient outcomes.
- Sepsis vs. septic shock. It is hypothesised that improvements in patient survival will be greater in patients with septic shock who receive prolonged infusion of beta-lactam antibiotics. Patients with septic shock commonly develop extreme pathophysiological changes, which may reduce effective % fT_>MIC_ beta-lactam exposures, and these patients are usually infected with pathogens that are less susceptible to antibiotic therapy (i.e., high MICs).
- Male vs. female sex. It is hypothesised that improvements in survival will be greater in male patients who received prolonged infusion of beta-lactam antibiotics. Critically ill male patients are more likely to demonstrate increased glomerular filtration rates leading to reduced % fT_>MIC_ betalactam antibiotic exposures. As beta-lactam antibiotics are predominantly cleared via renal elimination, prolonged infusion dosing may confer clinical advantages by maintaining effective beta-lactam antibiotic exposures throughout the dosing interval when compared with intermittent infusion.

The credibility of any subgroup analysis will be assessed using the Instrument for assessing the Credibility of Effect Modification Analyses (ICEMAN) in meta-analyses of RCTs (50).

### Assessment of evidence

The Grading of Recommendations Assessment, Development, and Evaluation (GRADE) approach will be used to evaluate the overall quality of evidence for each outcome measures (51, 52). Findings will be presented in a “Summary of findings and certainty of evidence” table. The certainty of evidence will be assessed based on five domains including the risk of bias, imprecision, inconsistency, indirectness and publication bias. For each outcome, the quality and certainty of evidence will be rated as “high”, “moderate”, “low” or “very low”.

### Patient and public involvement

As this is a secondary analysis, patient or consumer representation was not involved in the development of this protocol.

## Data Availability

Data sharing requests will be handled in accordance with The George Institute for Global Health (TGI) data sharing policy (https://www.georgeinstitute.org.au/data-sharing-policy).

## Ethics and dissemination

Human research ethics approval is not required as the study involves the use of existing collections of data that are de-identified. It is expected that the findings of this systematic review/meta-analysis will be presented at national and international intensive care and infectious diseases meetings. The results will be published in a peer-reviewed journal in the intensive care or infectious diseases literature. The publication will be made available on publicly accessible institutional websites. The results will not be publicly released until the main studies are published and are publicly available. Data sharing requests will be handled in accordance with The George Institute for Global Health (TGI) data sharing policy (https://www.georgeinstitute.org.au/data-sharing-policy).

## Discussion and limitation

This systematic review and meta-analysis will provide the most robust and up-to-date evidence concerning the clinical benefits of prolonged infusion versus intermittent infusion dosing of beta-lactam antibiotics in critically ill patients with sepsis or septic shock. New combined data from two large multicentre RCTs will be included to help address the uncertainty in beta-lactam antibiotic dosing strategy for ICU patients with sepsis or septic shock.

We acknowledge that there will be limitations in this systematic review and meta-analysis due to studies with heterogeneous ICU patient populations, variable illness severity, variable beta-lactam antibiotic dosing regimens, and differences in primary and secondary outcomes definitions.

